# Communication interventions to promote vaccination uptake: A systematic review protocol

**DOI:** 10.1101/2022.08.25.22279223

**Authors:** Daniella Castro-Barbudo, Oscar Franco-Suárez, Nathaly Aya Pastrana, Sandra Agudelo-Londoño, Deivis Nicolas Guzman-Tordecilla, Vidhi Maniar, Andres Vecino-Ortiz

## Abstract

**Background:** The use of communication interventions to promote vaccination has been more frequent in this century. These types of interventions have proven to be effective in reaching the majority of the population. Knowing the characteristics and results of communication interventions to foster vaccine uptake is important, especially with the outbreak of new infectious diseases such as the novel coronavirus (SARS CoV-2). This protocol will guide the development of a systematic review of the literature aiming to identify and analyze the evidence of communication interventions to incentivize vaccine uptake among populations living in low and middle-income countries (LMICs).

**Methods:** This systematic review protocol is guided by the Preferred Reporting Items for Systematic Reviews and Meta-Analysis Protocols (PRISMA-P). The searches for the systematic review will be carried out through five electronic databases PubMed, Scopus, Web of Science, Redalyc and PyscInfo. Two individuals will review each paper individually and in parallel using the software Rayyan. Duplicate elimination, title and abstract screening, and full text screening will be performed by the two reviewers. A matrix constructed in Excel will be used to extract data and to review the quality of the studies Quality assessment will be conducted using the QATSDD Critical Appraisal Tool.

**Discussion:** The results of this systematic review will contribute to the public health literature in the area of behavior change communication in general, and vaccination communication in particular. Findings from this study would also inform the development of communication interventions to improve vaccination uptake in LMICs.

**Systematic review registration:** PROSPERO CRD42022355541

## INTRODUCTION

Infectious disease outbreaks, especially those of respiratory transmission, have been a great challenge for public health and governmental management. In the search for the best strategies to protect populations from these diseases, public health has a cost-effective and rapid strategy in the process of containing the spread of respiratory transmission events: this is the case of vaccination (Berdasquera, Cruz, & Suárez, 2000). The success of this strategy depends on broad coverage and equity in its implementation (Casserly, 2005; (Becerra & Mújica, 2016). For this reason, a central concern for public health has been how to maintain vaccination coverage at high rates and how to guarantee immunization of the most at-risk groups and overcome barriers to access to health services. (De la Hoz-Restrepo, 2021).

The recent health crisis caused by COVID-19 has revealed many shortcomings at the state level with respect to health, which hinders equity and coverage in the vaccination process, leaving vulnerable populations unprotected. For this reason, it is necessary to think and build other forms of intervention to improve communication between health systems and the population. Some studies suggest that a communicative approach in health interventions can reduce hesitancy to get vaccinated or help to localize contagion hotspots and build better intervention strategies. (Lawes-Wickwar, S.; Ghio, D., et al, 2021; De la Hoz- Restrepo, 2021).

Communication interventions play a fundamental role in people’s decisions to adopt certain healthy behaviors. With the advent of new technologies and communication tools, many messages have been adapted to the particular needs of specific groups or individual users, and their low cost also makes them a better and more effective strategy for health systems. (Peter, C. et al, 2014; Gerend, MA. Et al, 2007) For this reason, “more high-quality research evaluating the impact of vaccine messaging on behavior outcomes is needed for firm conclusions to be made” (Lawes-Wickwar, S.; Ghio, D.; et al, 2021). Seeking to contribute to this, this protocol will guide a systematic review aiming to synthesize evidence of vaccine uptake communication interventions in low- and middle-income countries (LMICs). For this purpose, a brief literature review of the conceptual framework guiding the search and the methodological aspects considered in the search for quantitative and qualitative evidence, as well as for its analysis.

## CONCEPTUAL FRAMEWORK

This review is guided by various communication empirical and theoretical developments, two are underlined. The first is associated with health promotion; according to the WHO, this concept is defined as “the process of enabling people to increase control over, and to improve, their health. It moves beyond a focus on individual behavior towards a wide range of social and environmental interventions.” This process allows governments and communities to design and promote public health strategies to cope with health crises and challenges. It also has been proven to be an effective tool to public policy design in public and global health, leading to beneficial health outcomes (kumar y Preetha, 2012). Related to the above, Health Communication is a framework built to protect and promote the health of individuals, communities, and nations by influencing their decisions to have positive health outcomes. Thus, health communication framework focuses on providing “information, advice, and guidance to decision-makers (…) to prompt action that will protect [health]” (WHO, 2017)

The concept of Social and Behavior Change Communication (SBCC) was also considered in this review protocol. Various definitions of SBCC exists, one of them used by the United States Agency for Development Cooperation (USAID) defines SBCC as “the systematic application of theory-based, research-driven communication strategies to address individual level change and change within broader environmental and structural levels” (USAID, 2019). SBCC is usually used to influence certain behaviors at individual and community level. These strategies have been extensively used to promote prevention measures in communicable diseases, such as HIV (Bose et. al., 2022).

For this review protocol, communication interventions are seen as all message delivery strategies sent through different channels that aim to influence behavior change. Within the health field, these interventions are aimed to change behaviors that have a direct impact on people’s wellbeing and life quality. Regarding this, Yu says: “An important goal for health communication scholars is to test and evaluate the message strategies that can most effectively engage the target audience, raise their awareness of many health issues, and ultimately change their behaviors.” (Yu, N; Shen, F, 2013) The evidence indicates that behavior influences were enhanced when brief, risk-reducing or relative risk-framing messages were delivered; emphasized the benefits of vaccination to society; and addressed capacity beliefs and concerns among target populations. Thus, messages that were clear, credible, and in language that target groups could understand were associated with greater acceptability (Lawes-Wickwar, S.; Ghio, D.; et al, 2021).

Thus, many health outcomes regarding vaccines have shown to be improved by communication interventions, as changes in acceptability are mediated by psychological processes, and public health campaigns should formally consider variables such as intentions, beliefs, and gaps in understanding of vaccines and how they work. (Lawes-Wickwar, S.; Ghio, D.; et al, 2021). There are many factors involved in the acceptance of vaccination, such as inadequate and misleading information leading to a poor perception of vaccine safety. Therefore, vaccine uptake will be understood as a process influenced by social processes that shape perceptions and that can be influenced by communication strategies that seek to foster people’s interest and voluntariness and stimulate actions to increase vaccination. (Carter, J. et al, 2022)

## OBJECTIVES

### Review question

How have immunization communication interventions been designed and implemented in low- and middle-income countries and what results on health have been reported?

### Secondary questions

1. What communication interventions have been designed to promote/motivate vaccination uptake? (Who designed it, on which audiences was the interventions focused? Disease addressed, description of the intervention)
2. What theories, approaches and/or methodologies guided the development of the messages?
3. What messages, what type of messages, how often, through which communication channels, by whom, and to which audiences have the immunization communication interventions been implemented?
4. What were the reported outcomes of the interventions examined?
5. What methodologies and measurements were used to evaluate the results of the interventions?
6. How have the interventions considered intersecting axis of inequalities? (e.g., gender, ethnicity, social class)

## METHODOLOGY

To examine the scientific evidence about communication interventions to promote population behavior change about vaccination, this protocol follows the guidelines for Preferred Reporting Items for Systematic Reviews and Meta-Analysis Protocols (PRISMA-P) (Shamseer et. Al., 2015; Page, et al., 2021).

### Study selection criteria

Qualitative and quantitative articles that meet the selection criteria and are indexed in any of the selected databases will be included. Likewise, studies that have been published in journals with peer-review publication processes will be included. Only papers published in Spanish, Portuguese or English will be included. There will be no restrictions by the year of publication; hence no time limits are considered as exclusion criteria. The inclusion criteria will be described below using the PICO strategy as a reference:

– <u>Population:</u> no sex, age, or specific clinical conditions will be considered as exclusion criteria. Only we include LMICs studies.
– <u>Intervention:</u> Communication interventions (such as Interactive Voice Response (IVR) systems, text messages, social network messages, among others); behavior change interventions (such as social marketing, communication for behavioral impact (COMBI), social and behavior change communication (SBCC), communication for development (C4D), mhealth, health promotion, health communication, health economics. The search will focus on interventions focused on influencing the behavior of individuals and communities.
– <u>Comparison:</u> No comparison considered.
– <u>Outcomes:</u> Vaccination uptake influences. (Elements that influence vaccine uptake).

### Search methods

The search will be carried out through five electronic databases PubMed, Scopus, Web of Science, Redalyc and PyscInfo. Additionally, further articles will be obtained by reviewing the bibliographic references of each article reviewed. The search strategy in each database will follow the tools and requirements of each site. A pilot test of the search terms in each database will be carried out to identify the most efficient combinations of terms. Moreover, a pilot search on each database will also be conducted.

### Search Terms

The table below summarizes the search criteria for each database to be used:

**Table 1.**
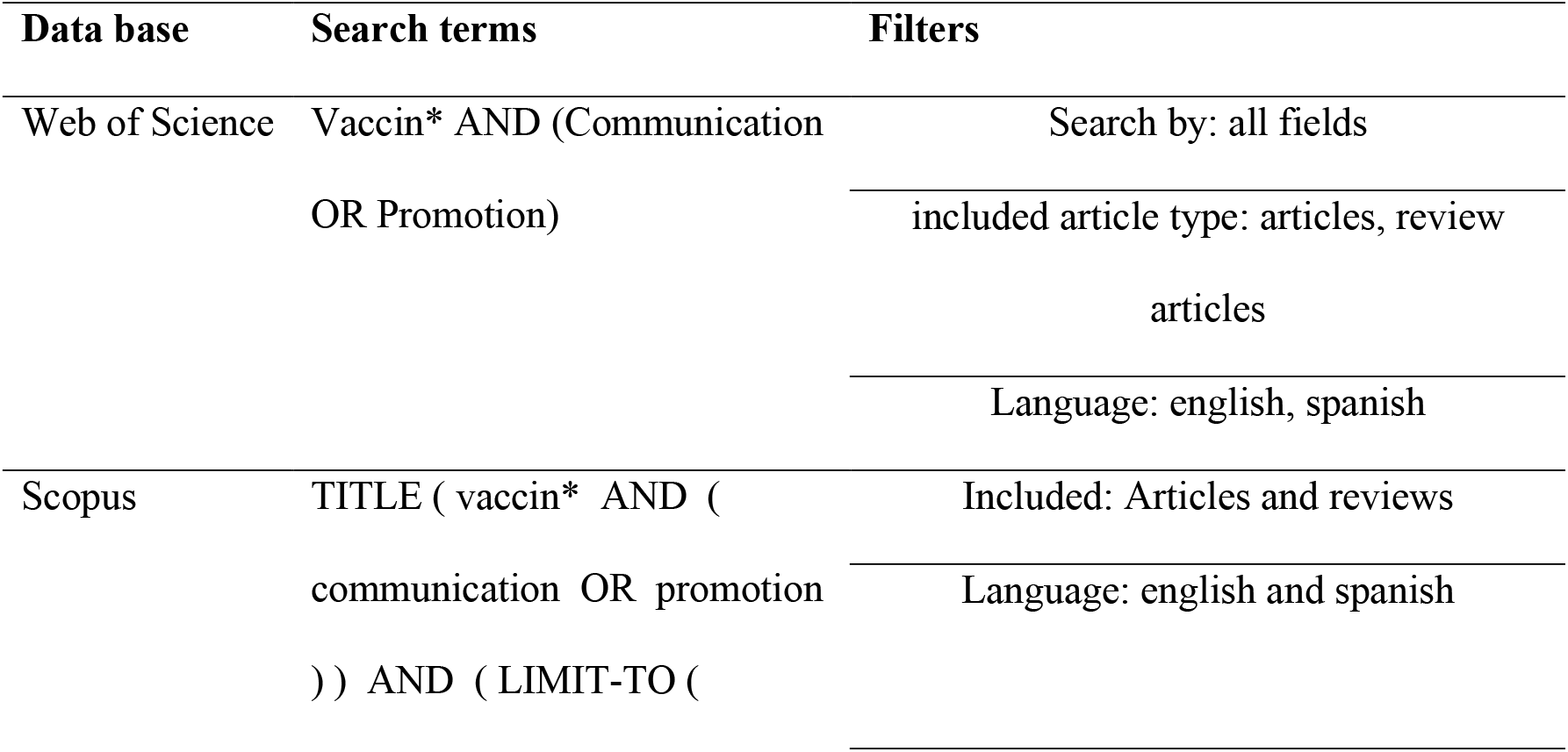
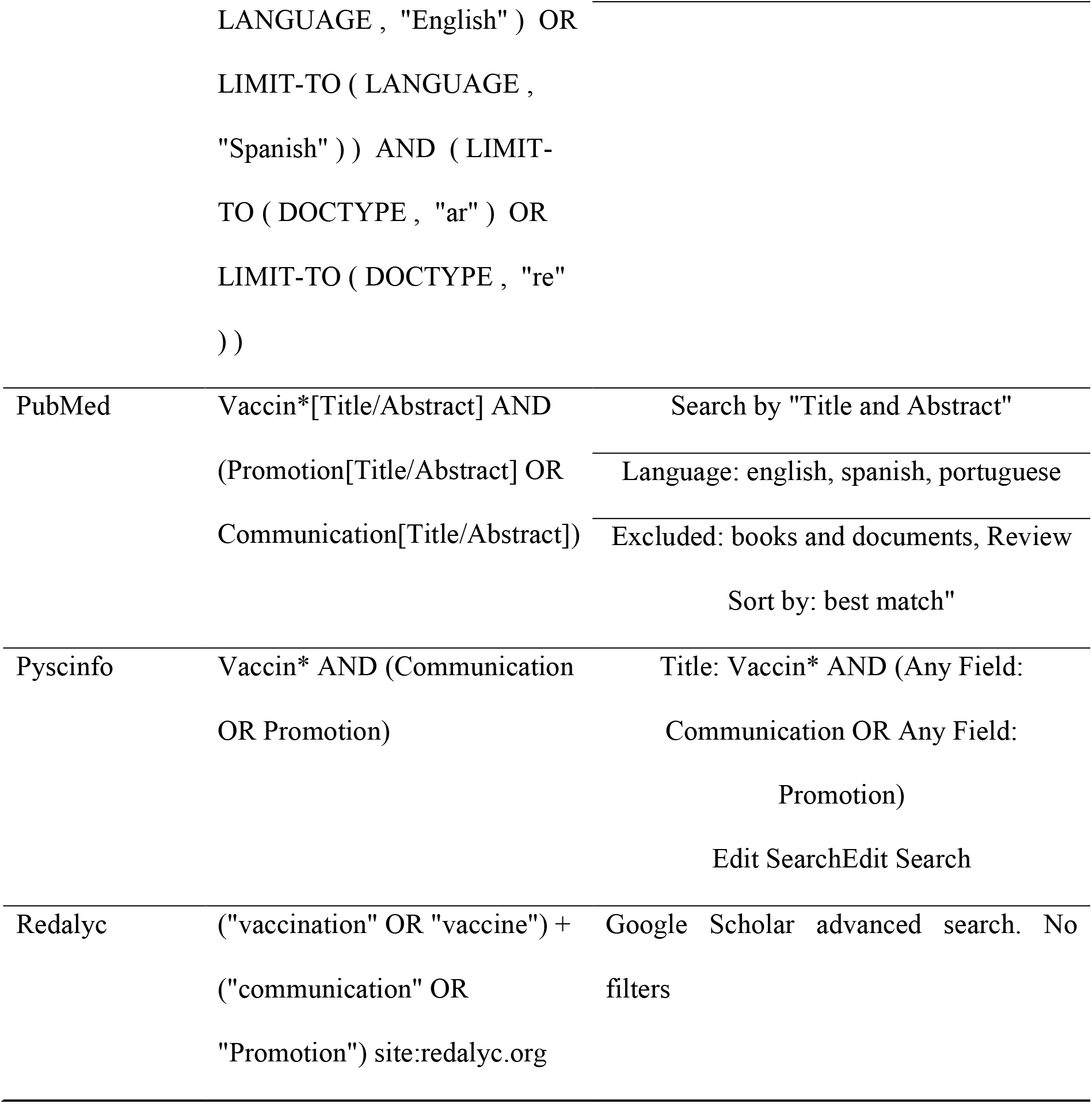
Search terms and filters used in each database

### Data management

The items obtained in this phase will be uploaded to the software Rayyan, which will facilitate the compilation and organization of all study records. The Rayyan software will be used to remove duplicate documents; however, we will continue to monitor this throughout the review to remove manually, if necessary, duplicate items that have not yet been removed.

### Selection process

#### Review of titles and abstracts

Once duplicates have been removed, reviewers will independently and thoroughly read each title and abstract of the articles uploaded into the Rayyan software and identify, (1) in the title the main topic of the study and the keywords included in the search of interventions and, (2) the PICO criteria for inclusion. The following types of publications will be excluded: proceedings or abstracts of a scientific event, letters to the editor, reviews, and editorial comments. Discrepancies between reviewers will be resolved by consensus; if no agreement is reached, a third reviewer will make the final decision.

#### Full text review

At this stage, the inclusion and exclusion criteria should be considered in greater detail. All articles that reach this phase will be saved in a shared folder, differentiating the excluded articles from those that were included for the final review. If necessary, the authors of the articles will be contacted to request additional information. Once the screening process is complete, a flow chart will be designed using PRISMA guidelines to summarize the screening.

### Data extraction

For data extraction, the complete documents will be saved in a shared folder. The reviewers will evaluate the texts independently and in parallel, they will use an Excel matrix that will be built and that will have all the information of interest for the necessary review. To ensure the quality of the extraction, a pilot will be made with one article. Some of the relevant information to be extracted from each document will be related to: Who designed the intervention, what was the objective of the intervention, what was the audience of focus, what were the characteristics of that audience, what theories or approaches were used, what are the characteristics of the messages used, what materials were used, how often was the message delivered, what were the health outcomes, among others.

### Risk of bias (quality) assessment

The reviewers will assess the methodological quality of the included studies and discrepancies will be discussed for consensus. In this quality assessment process, the QATSDD Critical Appraisal Tool (Sirriyeh et. al., 2012), which “evaluates the congruence, transparency, and organized reporting of research processes” (Fenton, Lauckner, & Gilbert, 2015) of studies of qualitative, quantitative, and mixed designs, will be used. The instrument contains 16 criteria, which have different scoring: from 0 to 3 (0 = not at all, 1 = very little, 2 = moderately, 3 = complete) (Aya Pastrana N et. Al., 2020). The results will be reported, and all studies will be included regardless of their quality score.^1^

### Strategy for data synthesis

To synthesize the findings a descriptive and narrative synthesis of the results will be made, addressing the advantages and disadvantages of interventions associated with the use of different type of messages to promote vaccination uptake in the population. All studies will be assessed for variability across populations, interventions, and outcome measures (we will describe the interventions and the populations in which they were tested, the limitations and goals met and the quality of studies) and the consistency of results. Tables will be used to present results.

### Meta-biases

This systematic review will not perform assessment of meta-biases within studies nor across studies.

## Discussion

The results of this review will be presented in one research article. The manuscript reporting the findings will be submitted to a relevant peer-reviewed journal for publication consideration.

## Data Availability

All data produced in the present work are contained in the manuscript

## List of abbreviations

LMICs: Low-and-middle-income countries
QATSDD: Quality Assessment Tool for Studies with Diverse Designs
COVID-19: CoronaVIrus Disease of 2019
WHO: World Health Organization
SBCC: Social and Behavior Change Communication
USAID: United States Agency for International Development
IVR: Interactive Voice Response
COMBI: Communication for behavioral impact
C4D: Communication for Development

## Declarations

### Ethics approval and consent to participate

Not applicable to this protocol.

### Consent for publication

Not applicable to this protocol

### Availability of data and materials

Not applicable to this protocol

### Author’s contributions

All authors conceived this review. DCB, OFS, NAP and SAL contributed in the development and refinement of the methodology. DCB and OFS developed the search strategy with the support of a librarian. DCB, OFS and NAP developed this manuscript, which has the approval of all authors.

## Acknowledgments

We thank Sandra Patricia Triana, librarian of Pontificia Universidad Javeriana and Sarah Elaraby, research of Hopkins University for their support providing feedback on our search strategy.

---

**Additional file 1:**
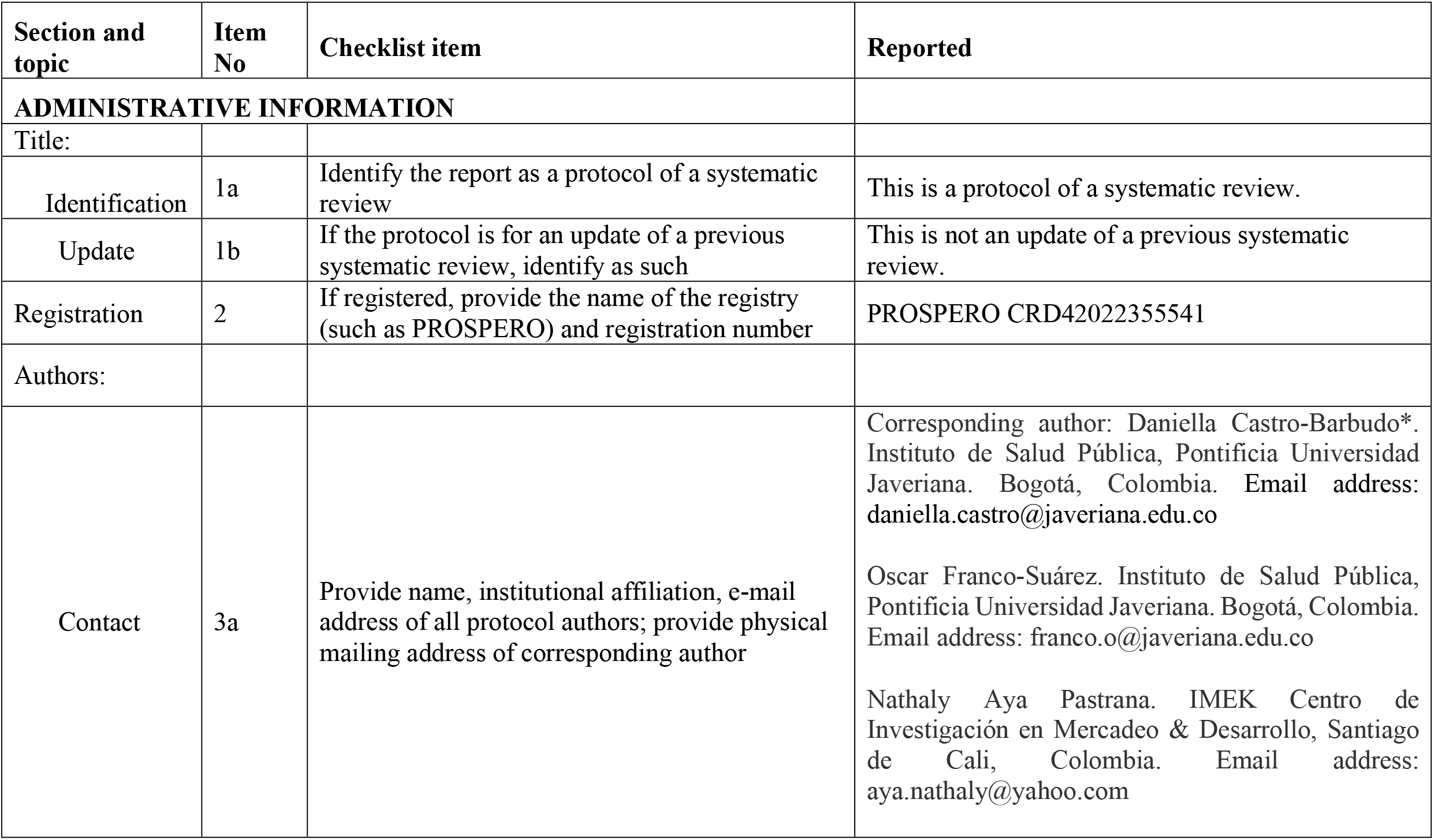
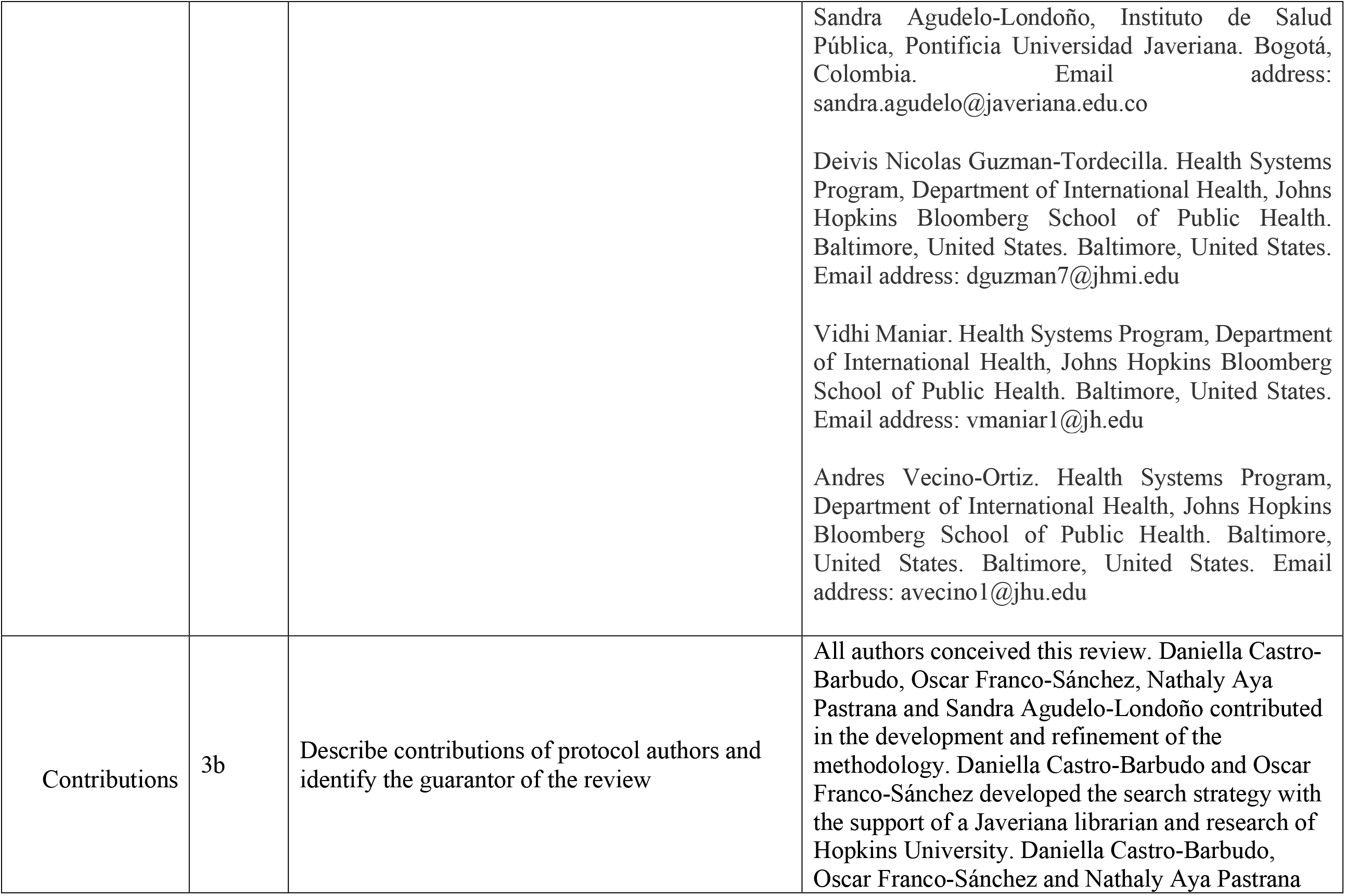
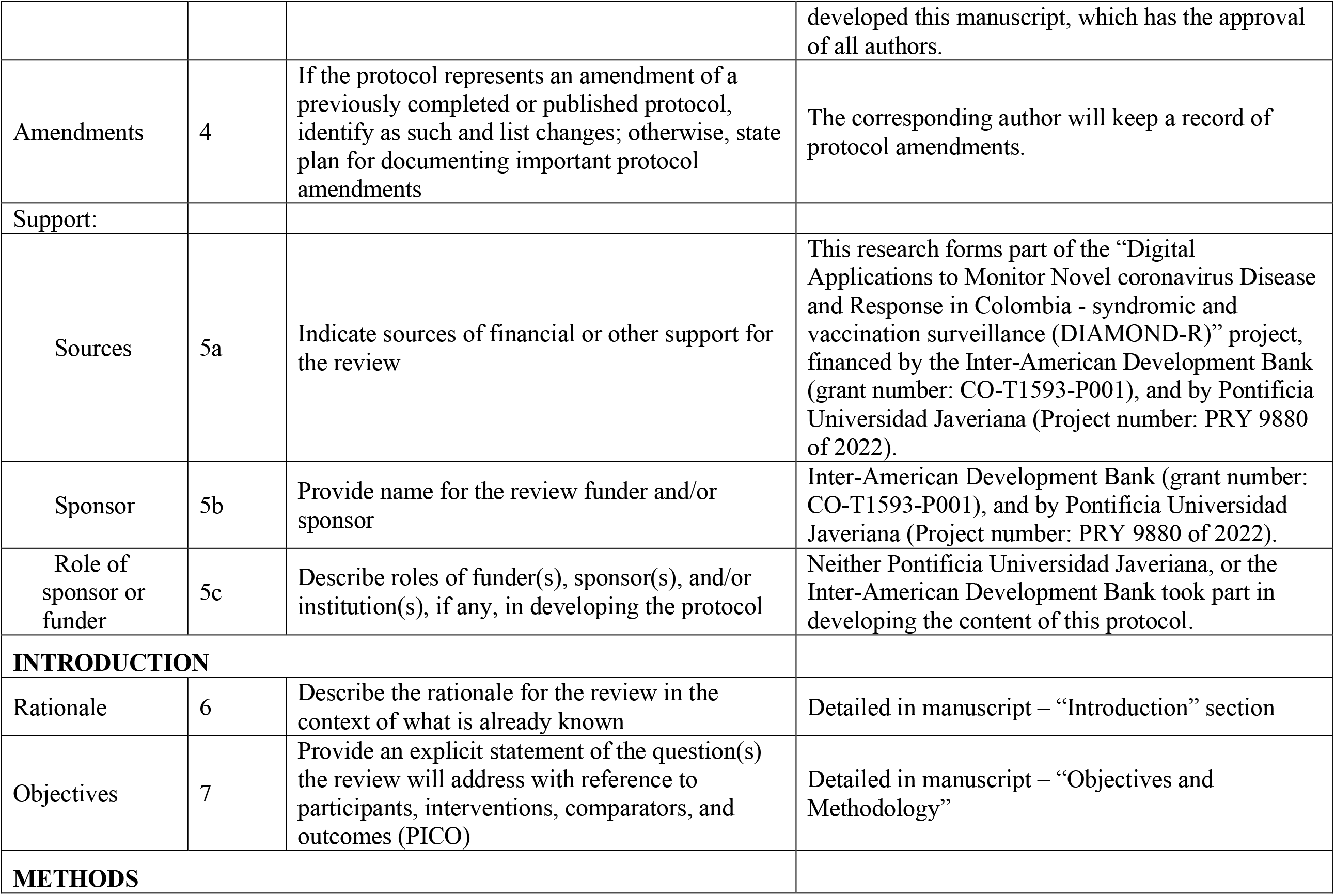
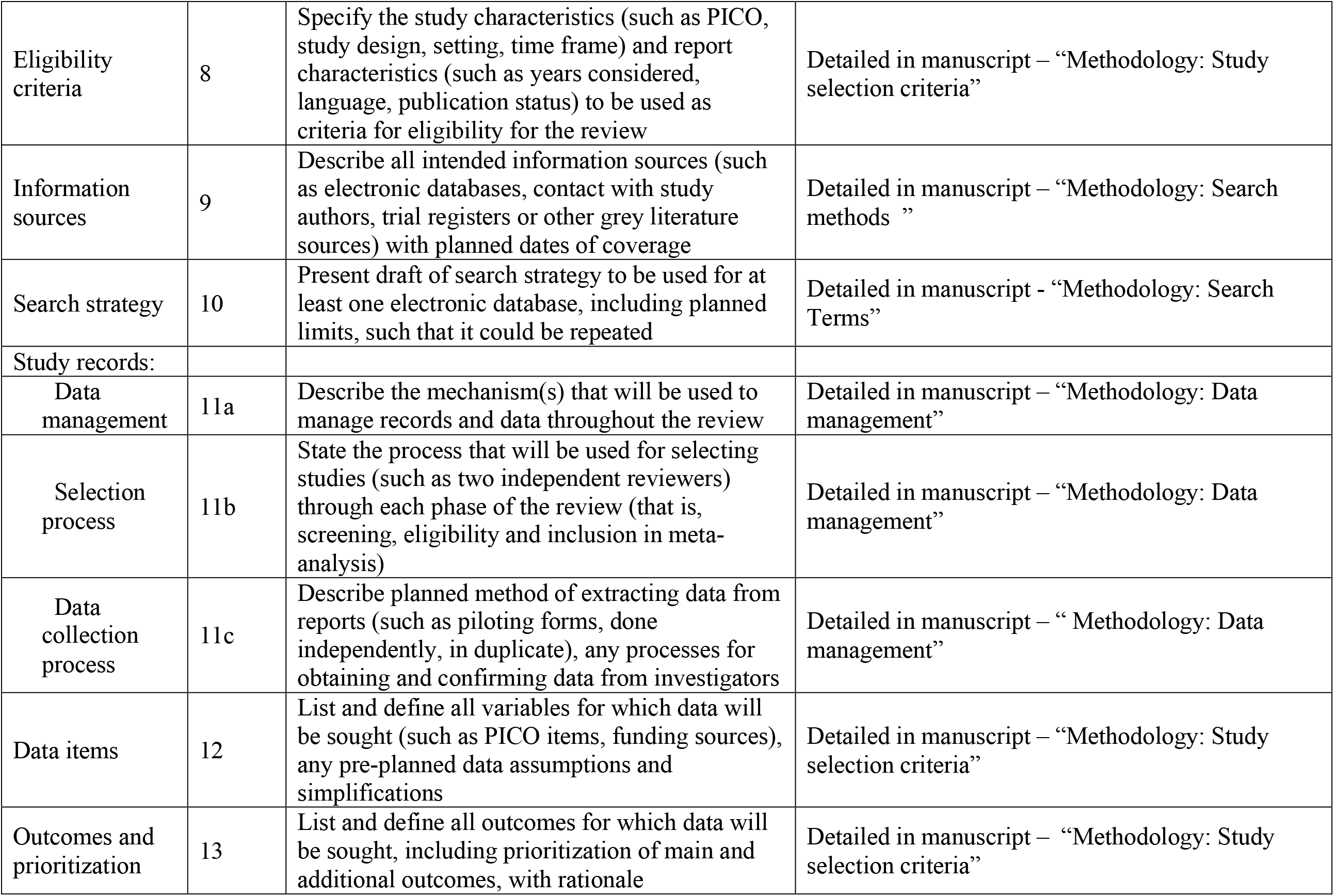
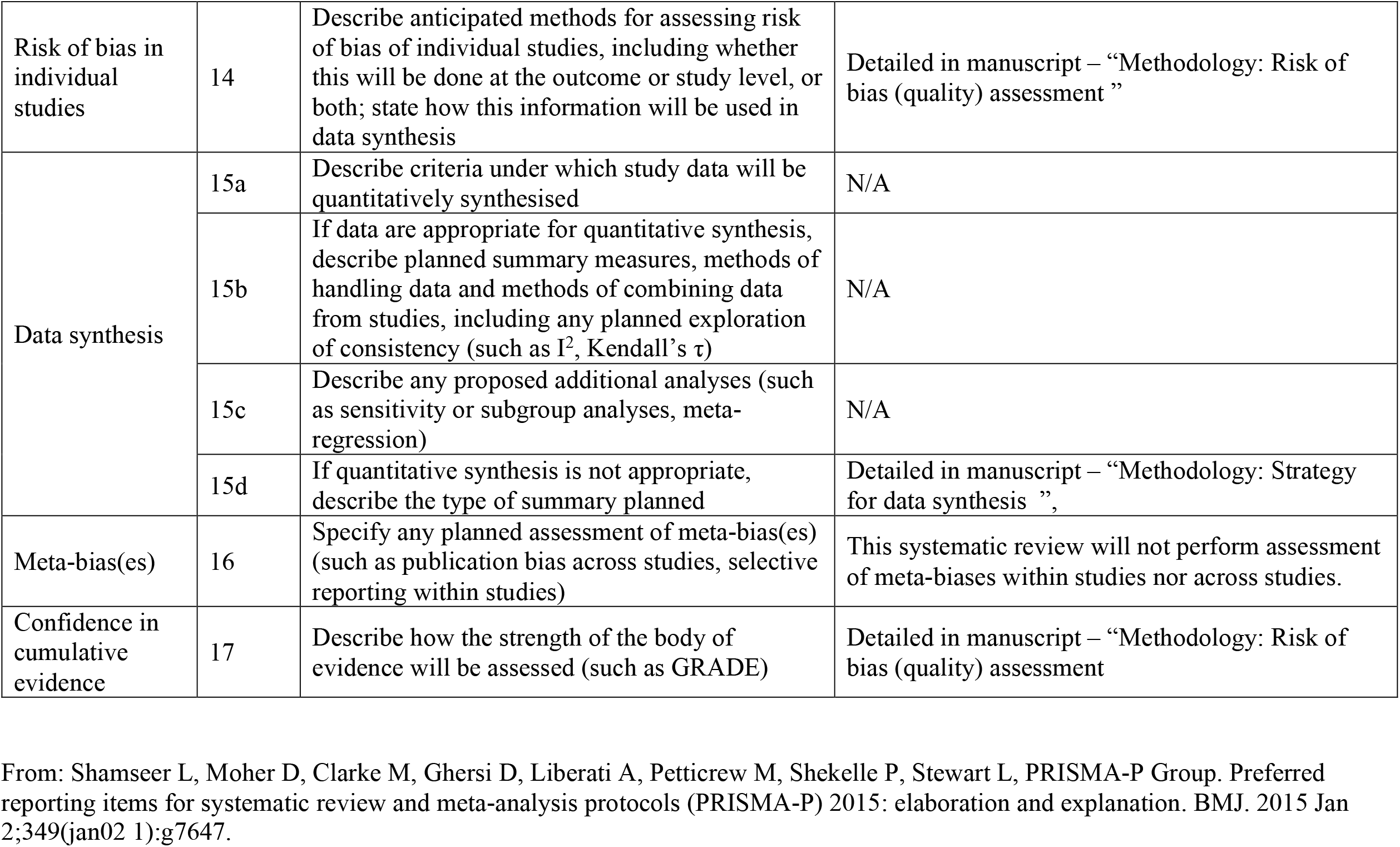
**PRISMA-P (Preferred Reporting Items for Systematic review and Meta-Analysis Protocols) 2015 checklist. Recommended items to address in a systematic review protocol**

## Footnotes

1 To see the tool used in the quality review, see Annex 2 attached to this document.

